# Microbial extract as a nature exposure material reduces the need for atopic dermatitis medication

**DOI:** 10.1101/2025.04.04.25325234

**Authors:** Johanna Kalmari, Iida Mäkelä, Vesa Hytönen, Heini Huhtala, Heikki Hyöty, Olli H Laitinen, Aki Sinkkonen

## Abstract

**Background:** According to the biodiversity hypothesis, interaction with microbially rich natural environments reduces the risk of immune-mediated diseases, such as atopy, allergies, and asthma. Urban environments have been associated with a higher prevalence of these diseases, and beneficial immunological and microbiological changes have been observed when contact with microbiologically diverse materials has been re-established in intervention studies. **Methods:** The current study is the first large, randomized, double-blinded, placebo-controlled clinical trial to test the effect of biodiversity intervention on atopic dermatitis. The study involved 142 atopic dermatitis patients divided into two groups: the experimental group used a topical lotion with an inactivated, diverse microbial extract as nature exposure material, and the control group used the same lotion without the microbial extract. The use of atopy medications was not restricted, but it was carefully recorded. Atopy severity was monitored using validated EASI, POEM and NRS itch scales. Skin condition was evaluated by TEWL and erythema measurements, and medicine use was recorded with questionnaires. **Results:** There were fewer atopy medication users and fewer atopy medication days in the experimental group compared to the control group, despite the similarity in eczema severity between the groups. Inactivated microbial extract prevented the deterioration of barrier integrity and the increase in erythema observed during the winter in the control group. There was no difference in adverse effects between the groups. **Conclusion:** Our results indicate that a skin lotion containing this type of nature exposure material could offer a feasible and safe way to support current treatments in atopic dermatitis.

## Introduction

Exposomic factors contribute to the development of various immune-mediated disorders such as asthma, allergies, and atopy ^1–5^. Exposome means all non-genetic exposures throughout the life span, such as living environment, climate, diet and exposure to environmental microbes ^6,7^. Urbanization and western lifestyle have been linked to a higher prevalence of atopy, allergy and asthma ^8–12^. Biodiversity hypothesis proposes that interaction with microbially diverse natural environment enriches human microbiome, enhances immune regulation, and provides protection against atopic and other immune-mediated diseases ^8,13–21^. Diminished contacts with diverse microbial communities may lead to the dysfunction of the immune system and finally diseases such as atopy and asthma ^8,14,22–24^. The significance of exposomic factors in immune-mediated diseases has been studied with populations sharing common genetic background, such as between Finland and Russian Karelia and Estonia ^8^. Urbanized Finland has more people sensitized to allergens compared to sparsely inhabited Russian Karelia ^8,25^. Microbial quantity and diversity are much greater in Russian house dust compared to Finnish house dust, and there is a dose-dependent association between the risk of atopy and the richness of microbial components in the dust ^8,25^. Cohort study from Finland and Estonia showed that the cover of forest and agricultural land within 2-5 km from the home was inversely associated with the atopic sensitization in children ^12^.

A farm environment in childhood reduces risk for allergic sensitization ^26,27^, and the high microbial diversity in rural environments that stimulates innate immunity has been proposed to be one protective factor ^8,14^. Instead, urban people are repeatedly exposed to human borne microbiota and less to diverse environmental microbiome from variable natural environments^8^. Consumption of drinking water with high microbial content has also been shown to be inversely associated with the prevalence of atopy ^11^. Also, high exposure to lipopolysaccharide (LPS) structures in gram-negative bacteria in the household has been recognized as protective against the development of atopy, but the benefit is affected by genotype ^28–30^.

Previous biodiversity intervention trials involving healthy adults and daycare children have demonstrated that exposure to microbiologically rich natural materials leads to beneficial microbiological and immunological outcomes. Specifically, forest-based soil and plant-based materials were shown to diversify children’s skin and gut microbiomes, reduce pathogen levels, increase regulatory T-cell (Treg) numbers, and shift the IL-10/IL-17 cytokine ratio in a more favorable direction ^18,19,31–33^. A placebo-controlled urban indoor gardening trial showed that daily exposure to microbiologically diverse soil enhanced immune regulation and skin microbiota ^34^. We have previously shown that use of a skin lotion containing diverse inactivated environmental microbiota was safe and did not lead to side effects in 12 patients with atopic dermatitis (AD) ^35^.

AD is a chronic and relapsing inflammatory skin disease, that is characterized by intense pruritus (itching), lesions, dryness and lichenification, with also other clinical features such as skin inflammation, barrier dysfunction and microbial dysbiosis with *Staphylococcus aureus* overgrowth ^36^. The treatment of AD focuses on immunosuppressive medication such as topical corticosteroids (TCS), tacrolimus and pimecrolimus or in severe cases biological drugs like dupilumab ^37–39^. AD patients and caregivers prefer to start with nonmedical treatments and move to corticosteroids and other medicines when AD symptoms become more severe ^40^. There is concern about the side effects of the medicines leading to a treatment strategy of using corticosteroids for as short a time as possible ^40^. Prolonged use of high-potency TCSs is potentially linked to adverse effect called topical steroid withdrawal (TSW). TSW or related topical steroid addiction (TSA) refers to physical dependence on medical corticosteroids with worsening of the symptoms after withdrawal of TCS use ^41,42^. Some AD patients also report decreased effectiveness of TCS after extended use ^42–44^. Due to lack of agreed diagnostic criteria and overlap of the symptoms with AD flares itself, the existence of TSW is still debated among medical community ^42^ although research evidence is growing ^45^.

Cutaneous bacteriotherapy approaches have been tested in AD including lysates of nonpathogenic bacteria, probiotics, bacteriophage-derived enzymes and skin bacterial transplants containing single or multiple coagulase-negative *Staphylococcus* strains ^46^. These therapies are based on selected, easy-to-grow bacteria of human origin, and the treatments aim at decreasing *S. aureus* colonization on atopic eczema and promoting skin microbial diversity ^46–48^. However, studies testing the efficacy of an exposure to highly diverse nature-derived microbial biodiversity in the treatment of AD have not been done.

The present study is the first randomized placebo-controlled double blind clinical trial where the efficacy of biodiversity intervention is studied among AD patients, most importantly as the need for AD medication and skin condition, analyzed as transepidermal water loss (TEWL) and erythema index. The principal idea was to bring environmental microbial diversity as an additive to normal life before the cold winter season begins that increases the risk for AD flares in Northern countries ^49^

## Materials and methods

### Experimental design

The study design was parallel group randomized, double blinded and placebo controlled. The study involved 142 Finnish voluntary adults suffering from AD (Figure 1). The regional medical research ethics committee of HUS (Hospital District of Helsinki and Uusimaa) gave an affirmative statement for this study (diary number 3842/2023). Written informed consent was provided by each trial participants before the study, and the trial has been registered in ClinicalTrials.gov (NCT06129136).

**Figure 1.**
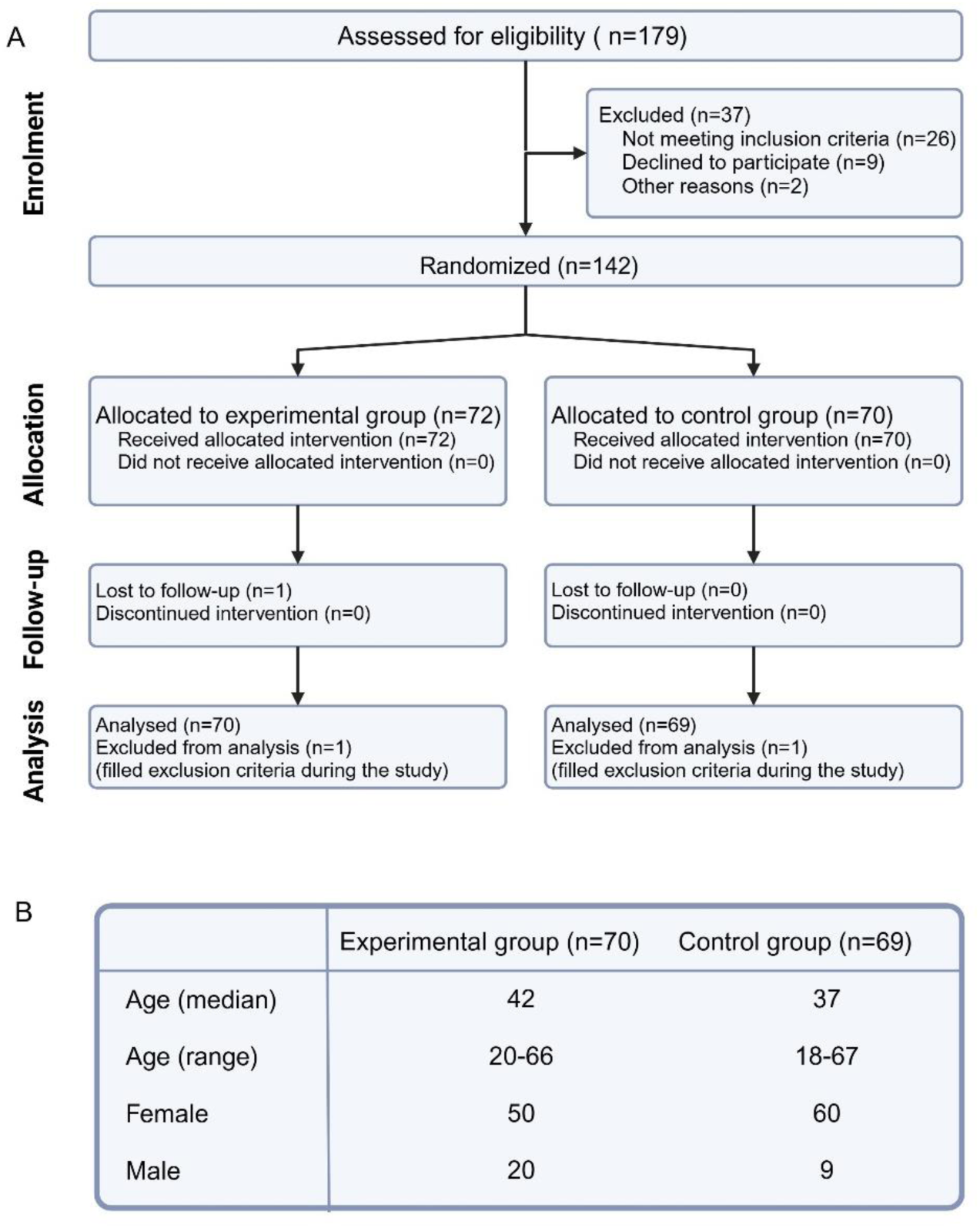
Study participant information. A) Flow diagram of the progress through the phases of parallel randomized trial of two groups, based on Schulz et al. 2010 ^51^. B) Baseline data of the trial participants. All data from clinical visits and questionnaires were analyzed, apart from the one lost during the follow up and two excluded before the analysis.

Trial participants were screened with an initial interview where the inclusion and exclusion criteria were reviewed. Inclusion criteria were fulfilling the Hanifin and Rajka criteria of AD ^50^ with AD eczema also in adulthood, age between 18-67, Finnish or Swedish language, and possibility to commit to the study protocol. Exclusion criteria were active cancer or cancer diagnosis within 6 months, immune suppressive medication (when not related to atopy or asthma), too severe AD (need for orally taken AD medicines other than antihistamines, or an injected AD drugs or clinical UV-light treatments within 6 months before the study or hospitalization because of AD two years prior to study), skin infection (such as an eczema that required antibiotic cream or a parasitic infection, e.g. scabies mite 6 months prior to study), psoriasis or other chronic skin disease besides AD, pregnancy or breastfeeding. Participants were equally randomized into two groups (72+70) by statistician outside the clinical team using a random number table.

### Description of participants

Altogether 142 participants started the study, one was lost during follow up and two were excluded because they filled the exclusion criteria, so the analyses included 139 (70 experimental and 69 control) participants (Figure 1). Fifteen subjects dropped out during the study (10.6 %): 8 in experimental (Exp) group and 7 in control (Ctrl) group. In addition, there were 7 participants (4 in Exp-group, 3 in Ctrl-group) that answered the questionnaires until the end, but who did not attend the final clinical visit. Their data were included in the analyses.

There were 19 participants (8 in Exp group and 11 in Ctrl group) that categorially declined to take any medicines except antihistamines to treat their atopy. Their refusal to use atopy medicines included corticosteroids and other medical creams, such as Tacrolimus and Pimecrolimus. These participants that followed TSW represent 13.7 % of all participants, and they are included in all other analyses except the analysis of medicine used to eliminate any bias resulting from categorical refusals of medication.

The experimental group used a skin lotion that contained 1% of inactivated microbial extract. The lotion was manufactured by Orion Pharma, brand name Aqualan L (INCI: Aqua, Caprylic/Capric Triglyceride, Isopropyl Palmitate, Glyceryl Stearate, Glycerin, Cetyl Alcohol, PEG-30 Stearate, Ceteareth-20, Ethylhexylglycerin, Phenoxyethanol). The microbial extract was manufactured and mixed with the lotion by Uute Scientific Oy, Finland. The extract was made from Finnish plant and soil materials, and it includes over 600 different bacterial species (Operational Taxonomic Units), their metabolic products as well as humic acids, mimicking Finnish forest soil biodiversity. Microbial extract was thermally inactivated with autoclaving (121°C, 2bar, 15min) before adding to the lotion. The same microbial extract and lotion mixture has previously been used in our previous safety study in the same 1 % concentration ^35^.

The control group used the same lotion Orion Pharma Aqualan L in which a biochemically neutral coloring ingredient was added (iron oxides, manufacturer Pure Rock Minerals Cosmetiques SL) to make it indistinguishable from the intervention material. Also this placebo product was the same as in our previous safety trial among AD patients ^35^. If the participants were not able to use Aqualan L due the incompatibility with their skin, an alternative product was skin oil Ceraderm by Decem Pharma Oy (INCI: Paraffinum liquidum, Isododecane, Isopropyl myristate, Simmondsia chinesis seed oil), supplemented with the inactivated microbial extract or with iron oxides. Lotions were made just before the trial started and new lotion batches were given at midpoint visit. One person outside the research team made the allocation to interventions and all other persons (trial participants, research team, data analyst) were blinded. Unblinding was performed after the database was locked and the result analysis phase was completed.

Participants came from various regions of Finland, mostly from the capital region Helsinki, but also for example from Tampere, Jyväskylä, Seinäjoki, Kuopio and Oulu (Oulu being the northmost region). Mean outside temperature in Helsinki and Jyväskylä areas (Fig 2A) were fetched from Finnish Meteorological Institute to indicate the weather conditions. This is relevant since the cold and dry air during the wintertime increases the incidence of flares in AD patients ^49^. In Figure 2A the temperature in August is a mean between the first study start date August 17^th^ and the end of the month and the temperature in May is a mean temperature calculated for the period between the 1^st^ of May and the last study date 17^th^ May.

**Figure 2.**
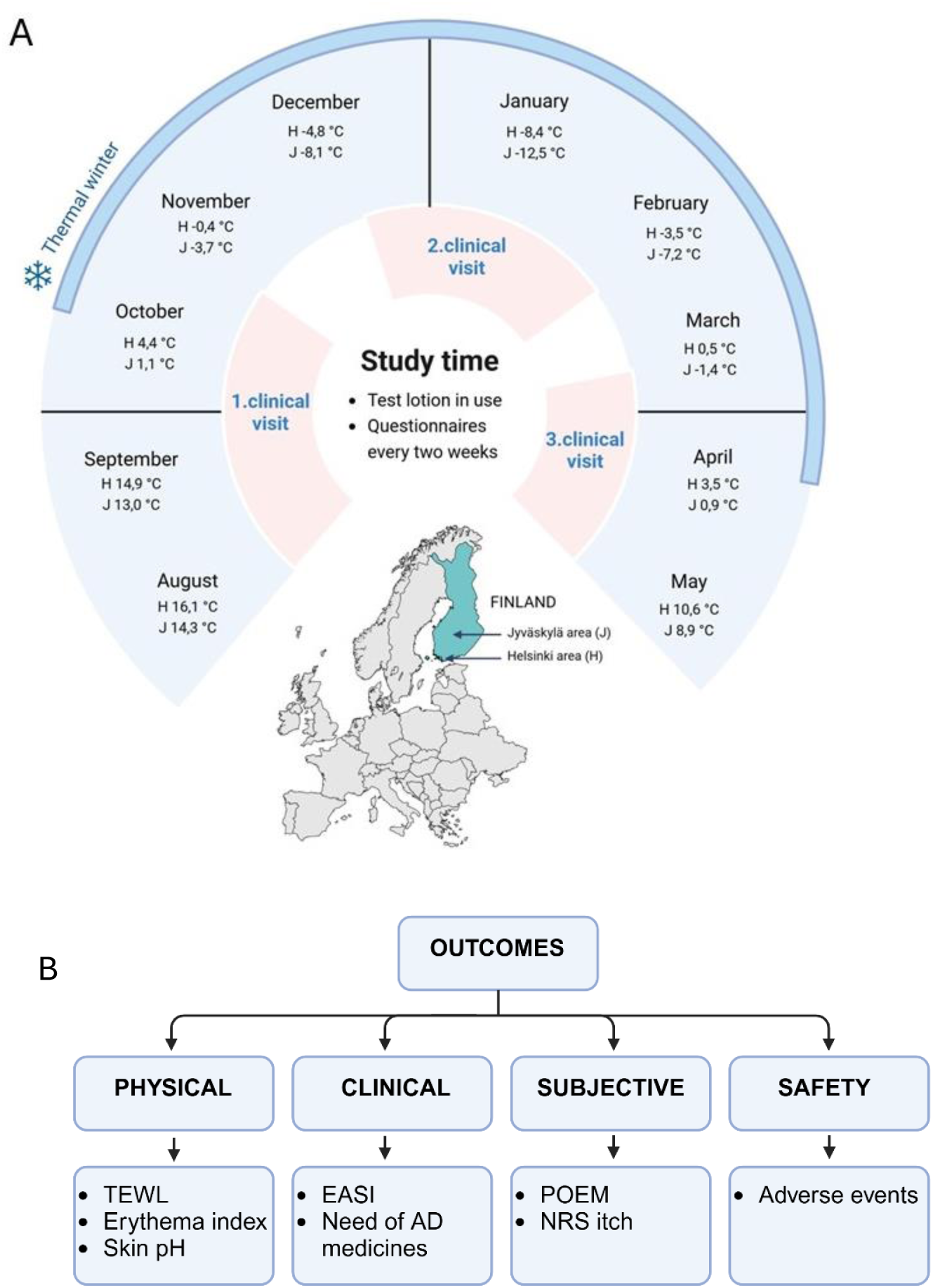
Design of the study. Schematic presentation of A) the study protocol (temperatures are mean values per month in Helsinki and Jyväskylä areas) and B) the measured variables. TEWL=Transepidermal water loss, EASI=Eczema Area Severity Index, POEM=Patient Oriented Eczema Measure, NRS itch=Numerical Rating Scale of itch.

### Research methods

Participants started to use the given test lotion after their first clinical visit, before the Finnish wintertime, and ended using the test lotion on the 3^rd^ visit, before the summer (Fig 2A). Each participant used their test lotion for 5-9 months, depending on when they arrived for the first and the last clinical visits.

The analytical methods and outcomes of the study are depicted in Fig 2B. Data on physical measurements (Transepidermal water loss (TEWL), erythema index and skin pH-value) and Eczema Area and Severity Index (EASI) scores were collected during the three clinical visits. Other data, need of AD and allergy medicines, Patient Oriented Eczema Measure (POEM), Numerical Rating Scale of itch (NRS itch) and data of adverse events were collected from on-line questionnaires (using Webropol -platform, Finland) filled every two weeks through the study period by the trial participants. There were no restrictions on the use of the medicines, but the need for AD medicines and antihistamines was monitored with questionaries.

There were two research skin areas for every trial participant: a standard research area just below the elbow bend of the dominant arm, and an eczema site, primarily selected also from arms (Fig 3). If there was no eczema or the eczema was too severe on the arms, then eczema research area was selected in co-operation with the participant from another body site. The standard research area was the same with every person, regardless of the existence of eczema in this area.

**Figure 3.**
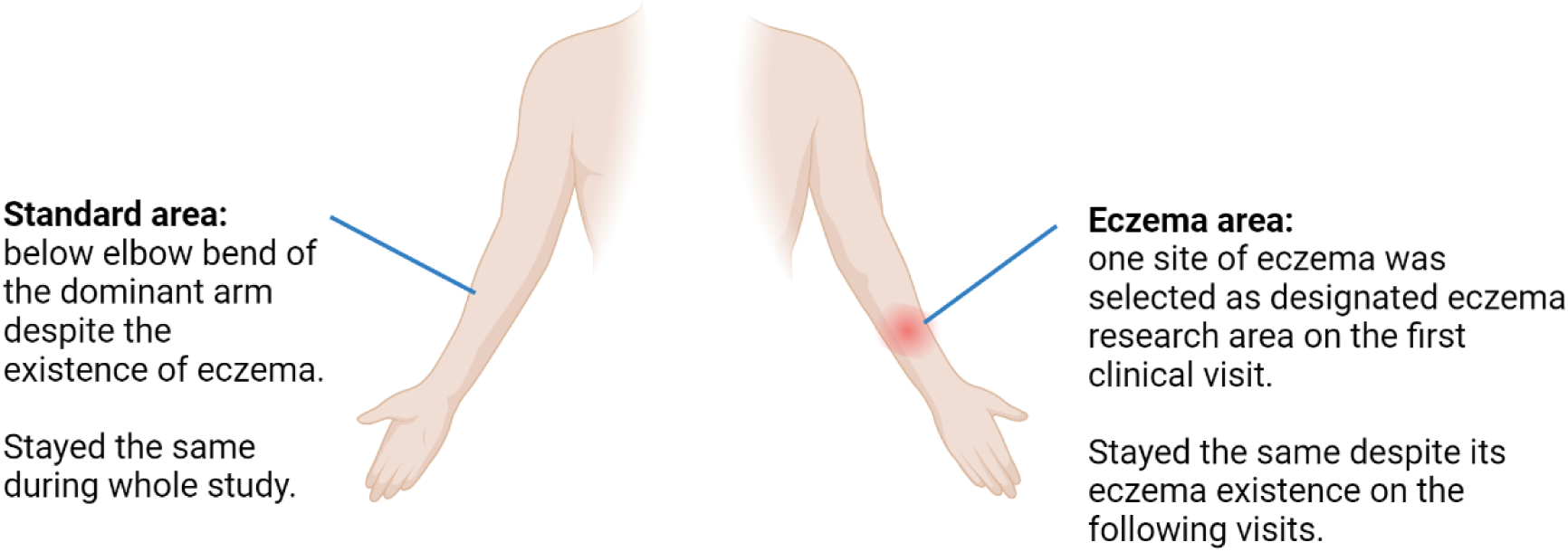
Designated research areas. The skin analyses were performed within designated areas as indicated in the drawing.

The test lotion was advised to be applied to at least to both arms and to the designated research areas at least three times per week. The measurements were taken from these designated research areas in all three visits. The research areas were kept the same during the whole study which was possible by taking photographs of the research areas in the first meeting and using these photos for locating the places in the next meetings. Photos of the research areas and the arms were taken in every meeting for follow up. Biorender was utilized for making the figures 1,2 and 3.

The validated EASI method has been discovered to be consistent over time, which is critical in the intervention study ^52^. EASI evaluation was done by the same trained investigator every time, following the instructions of Hanifin, J. M. *et al.* ^53^. EASI values range from 0 to 72, where 0 is clear, 0.1-1.0 almost clear, 1.1-7.0 mild, 7.1-21.0 moderate, 21.1-50.0 severe and 50.1-72.0 very severe ^53^.

The validated POEM method was stated as the core instrument for all AD clinical trials by the stakeholders of the fourth Harmonizing Outcome Measures for Eczema meeting ^54^. It is a method where patients self-evaluate the frequency of seven symptoms (itch, sleep disturbance, dryness, flaking, oozing, bleeding and cracking) in the past 7 days. POEM values range between 0 and 28 points, which are translated into disease severity as follows: 0-2 (clear/almost clear); 3-7 (mild); 8-16 (moderate); 17-24 (severe); 25-28 (very severe) ^55^.

As itch is a key symptom of AD, we included Peak Pruritus Numerical Rating Scale (later NRS itch) in the regular questionnaire. The question to be answered was “how would you rate your itch at the worst moment during the previous 24 hours?” on a scale from 0 to 10, with 0 being “no itch” and 10 being “worst itch imaginable”. NRS itch is regarded as reliable, sensitive and valid scale for evaluating worst itch intensity in adults ^56^.

### TEWL, Erythema index and pH measurements

Transepidermal water loss (“TEWL”) level was measured by Delfintech Vapometer ^57^ and Erythema index by Delfintech Skin Color Catch instrument. Skin pH value was measured using Skin & Scalp pH meter HI-981037 from Hanna Instruments. TEWL and Erythema results were calculated as mean of three repeated measures. Skin pH value was a single measurement due to long time needed for the value to settle.

### Statistics

Statistical analyses were done using IBM SPSS Statistics (version 29.0.1.0) and GraphPad Prism 9.0. The normality of the data was analyzed using Shapiro-Wilk normality test. For parametric data, the applied tests were Independent Samples t-test, Paired-Samples t-test and Pearson correlation. For nonparametric data, the applied tests were Mann-Whitney U-test for independent measures, Wilcoxon signed rank test for repeated measures and Spearman’s Rho to analyze correlations. Chi-Square Tests were applied for binary data. All p-values are two-tailed.

## Results

### Microbial extract applied on the skin decreases the use of atopy medication

We followed the usage of AD medication and observed fewer medication users in the experimental group (Fig 4A and B). The difference was particularly striking in the beginning of winter (the thermal winter started at the end of October Fig 2A). The number of medication users decreased in both groups during the study. The results were consistent regardless of whether the analyses were conducted chronologically based on the calendar (Fig 4A) or weeks from the start date of the trial for each participant (Fig 4B).

**Figure 4.**
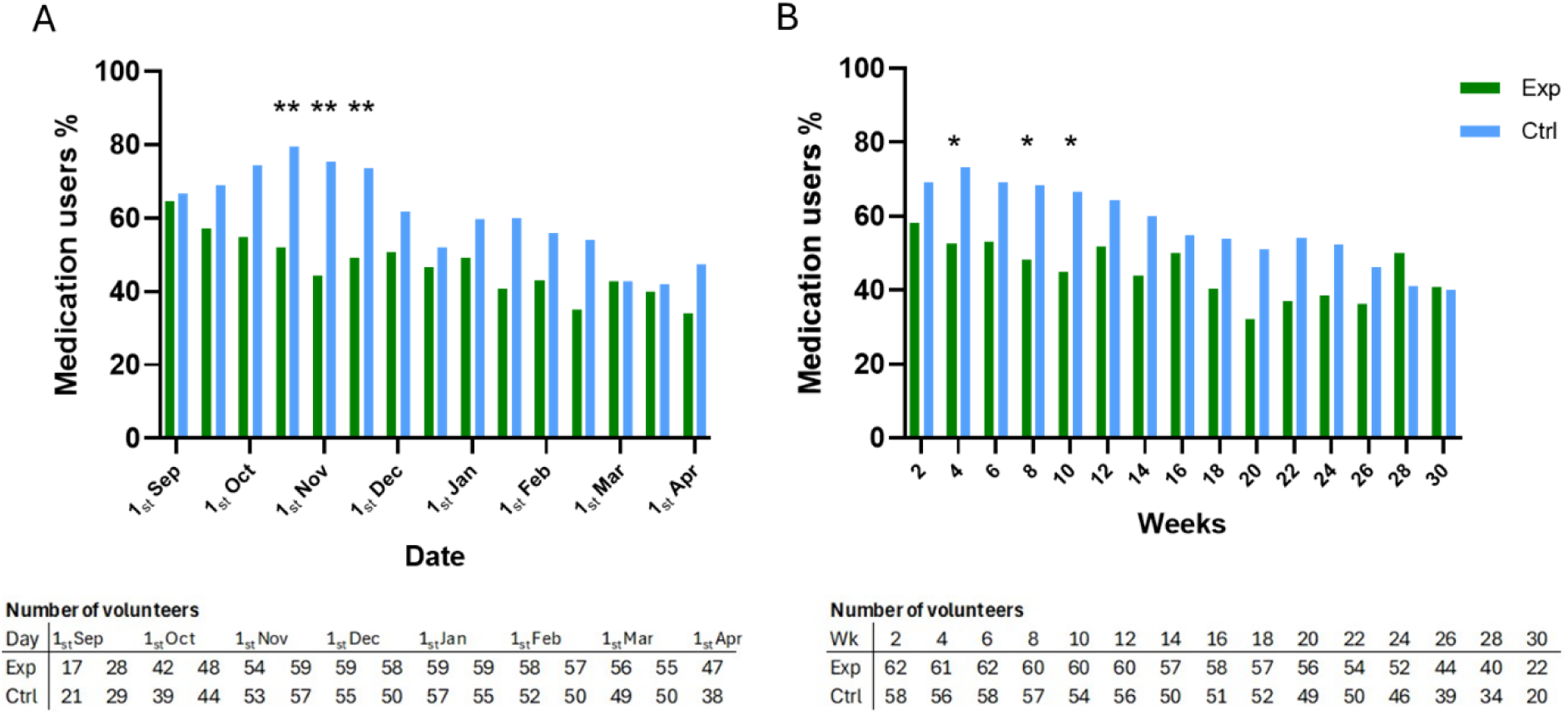
AD medication users during the study. The proportion of atopy medication users A) chronologically and B) in weeks from the start of the exposure in experimental (Exp) and control (Ctrl) groups and the number of answers gathered in each time point. * p ≤ 0.05, ** p ≤ 0.01.

When the proportion of medication users was analyzed in males and females separately, the reduction in difference in the use of medications was clear among women (Fig 5). In chronologically organized data (Fig 5A), p-values of the differences were 0.003 (15^th^ Oct), <0.001 (1^st^ Nov), 0.017 (15^th^ Nov), 0.005 (15^th^ Jan), 0.048 (1^st^ Feb). In data organized by study duration (Fig 5B), the p-values were 0.01 (wk 4), 0.038 (wk 6), 0.011 (wk 8), 0.018 (wk 10). On the contrary, there was no difference between the groups among men, just similar decreasing trend as in women, possible due to clearly lower number of men participants (Supplementary figure 1). There was no difference in the number of AD medication users between the groups during the three-month period before the study, either with all participants or when analyzed by gender (Supplementary figure 2).

**Figure 5.**
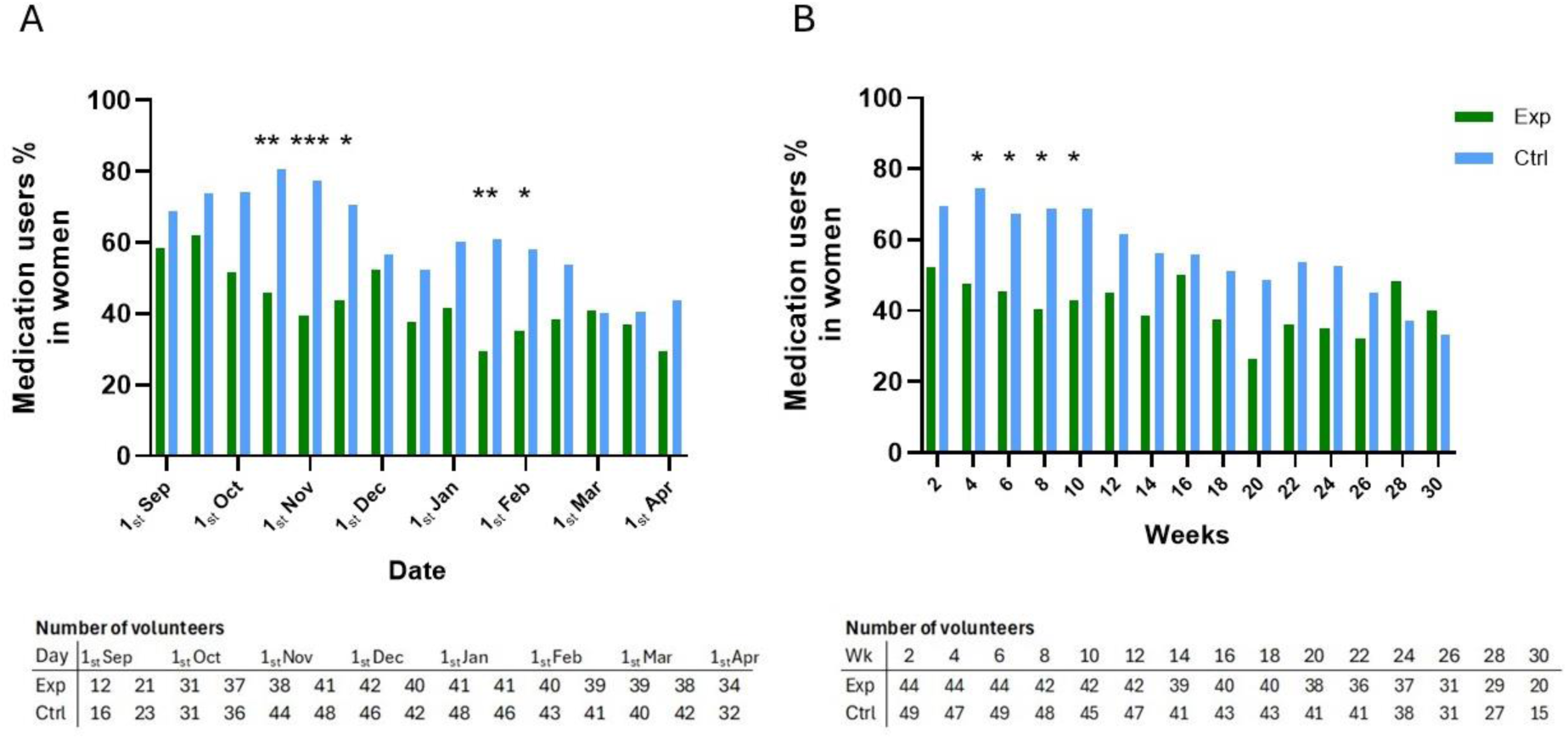
AD medication users by female participants. Atopy medication users among female participants A) chronologically and B) by weeks in experimental (Exp) and control groups (Ctrl) and the number of answers gathered in questionnaires at different time points. * p ≤ 0.05, ** p ≤ 0.01, *** p ≤ 0.001.

In addition to less atopy medication users (Fig 4, 5) there were also less atopy medication days in experimental group compared to control. That is seen in the proportion of medication days between the start and midpoint visits (Fig 6). The difference existed more precisely in the following time points: 1^st^ Nov (p=0.002), 15^th^ Nov (p=0.005) and 15^th^ Jan (p=0.042) (Supplementary Fig 3A), and at 4-week (p=0.024) and 10-week (p=0.015) questionnaires (Supplementary Fig 3B).

**Figure 6.**
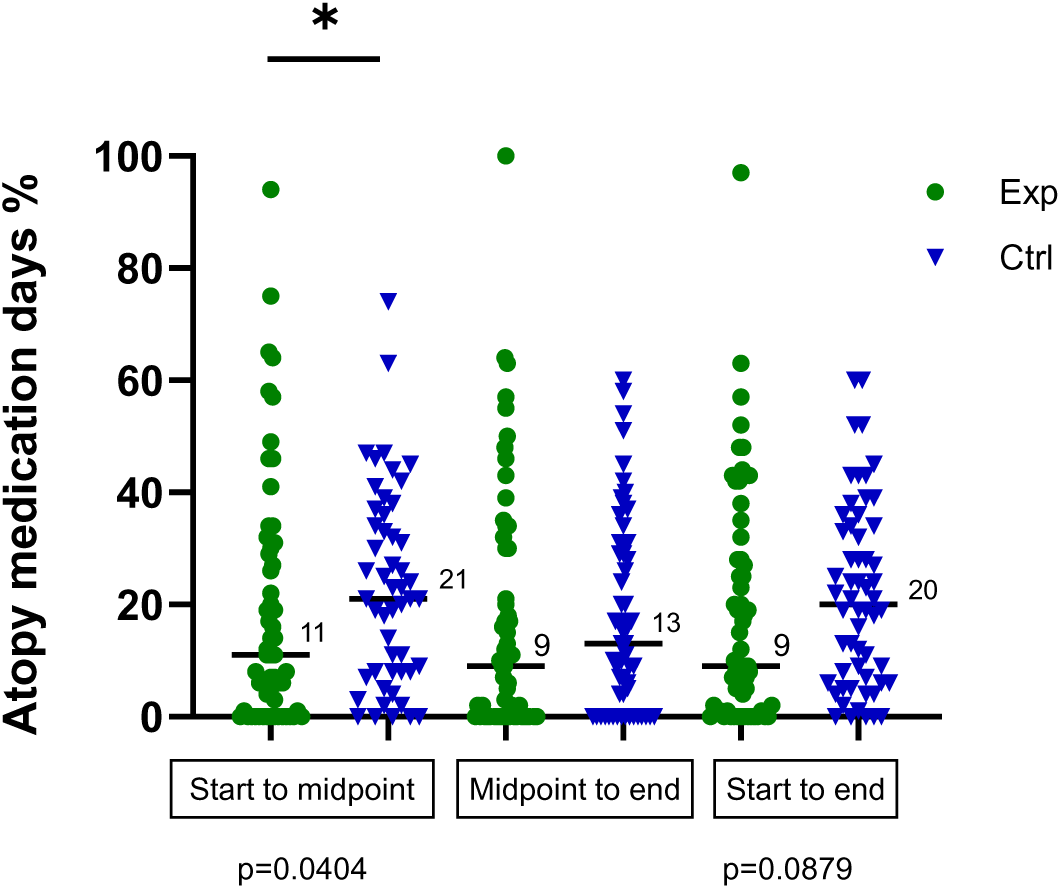
Proportion of AD medication days between the clinical visits. Lines with values represent medians. Exp=experimental, Ctrl=control.

The median of the longest medicine-free period during the study was 56 days in the experimental group and 42 days in the control group (Supplementary Fig 4). The difference between the groups was not statistically significant (p=0.1133), neither within women (p=0.0844) or men (p=0.731). There was no difference between the groups in terms of the total amount of topical AD medicines used in grams.

### There was no difference in eczema severity between the groups despite the difference in medication

Eczema Area Severity Index (EASI) decreased in both groups from start visit to end (p-values 0.0001 (Exp) and <0.0001 (Ctrl)) and from midpoint visit to end (p-values 0.0059 (Exp), 0.0008 (Ctrl)) (Fig 7A, descriptive statistics in supplementary table 1). Patient oriented eczema measure (POEM) showed a similar decreasing trend (Fig 7 B, C) as EASI and medication users during the study period (Fig 4B, C). There were no differences between the groups in EASI or POEM.

**Figure 7.**
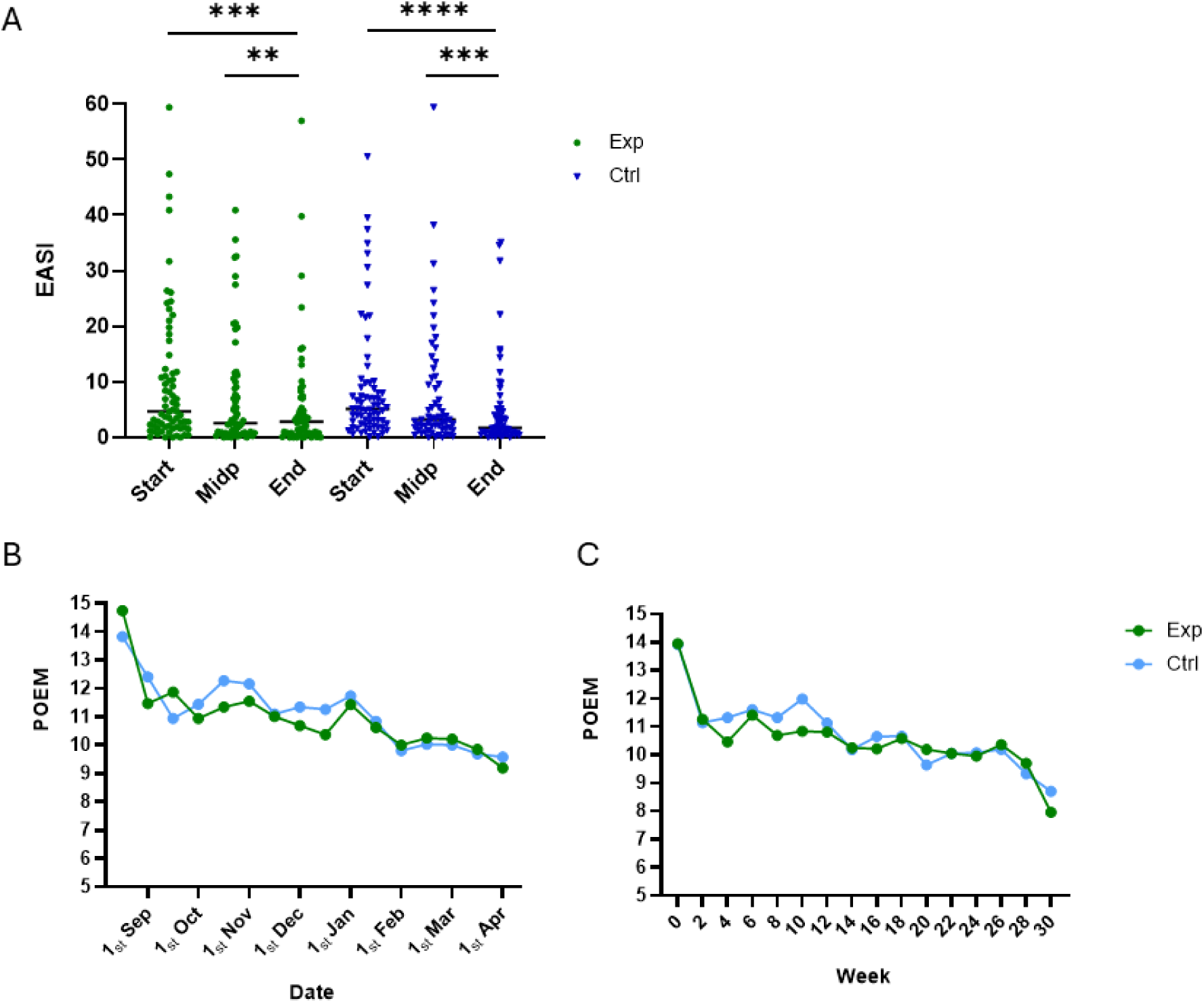
Eczema severity analyzed by clinician and by the trial participants themselves. A) Eczema area severity index (EASI) in the experimental (Exp) and control (Ctrl) groups in the three clinical visits during the study. Lines represent medians. B) C) Mean values of patient oriented eczema measure (POEM) in the biweekly questionnaires in chronologically (B) and weeks (C). * p ≤ 0.05, ** p ≤ 0.01, *** p ≤ 0.001, **** p ≤ 0.0001.

When the difference between the genders in EASI was analyzed, women had lower EASI at the midpoint compared to men (p=0.005). As there was no difference between the sexes in the beginning of the study, this finding underlines the overall benefit of the exposure (benefit in both groups) for women. There was no difference between the groups in women (midpoint p-value 0.069) or men EASI values.

Experimental group managed with less medication even though the eczema severity was similar with the control group (Table 1). The percentage of severe AD cases decreased in both groups during the study. EASI scores of dropouts were: 2 severe (1 exp, 1 ctrl), 4 (2 exp, 2 ctrl) moderate and 9 mild (5 exp, 4 ctrl), and obtained results are included into analyses until the discontinuation.

**Table 1.** Eczema severity in the experimental (exp) and control (ctrl) groups during the study. Percentages for each severity category are calculated based on the group size in each time point.

| Number of participants | EASI1 (baseline) |  |  | EASI2 (midpoint) |  |  | EASI3 (end) |  |  |
| --- | --- | --- | --- | --- | --- | --- | --- | --- | --- |
|  | Mild | Moderate | Severe | Mild | Moderate | Severe | Mild | Moderate | Severe |
| Exp | 42 | 17 | 11 | 43 | 15 | 6 | 46 | 10 | 4 |
| Ctrl | 45 | 14 | 10 | 43 | 12 | 6 | 47 | 8 | 4 |
| Exp % | 60.0 | 24.3 | 15.7 | 67.2 | 23.4 | 9.4 | 76.7 | 16.7 | 6.7 |
| Ctrl % | 65.2 | 20.3 | 14.5 | 70.5 | 19.7 | 9.8 | 79.7 | 13.6 | 6.8 |

### Lotion with the microbial extract prevented deterioration of the skin barrier in winter

The integrity of the skin barrier was evaluated by using TEWL measurement. At the eczema research area the difference between the groups at the start was statistically significant (Fig 8A, p=0.0308), but further analysis showed that there were no differences between the groups in fold changes (calculated by dividing the subsequent value with the original value, p-values were 0.950 from start to midpoint, 0.359 from midpoint to end and 0.457 from start to end) (Supplementary Fig 5A). There was a decrease in eczema area TEWL values in the control group during the study (p-values were 0.0133 from midpoint to end and 0.0022 from start to end) and a decreasing trend in the experimental group (p-value 0.0551 from start to end) (Fig 8A, descriptive statistics in supplementary table 2A).

**Figure 8.**
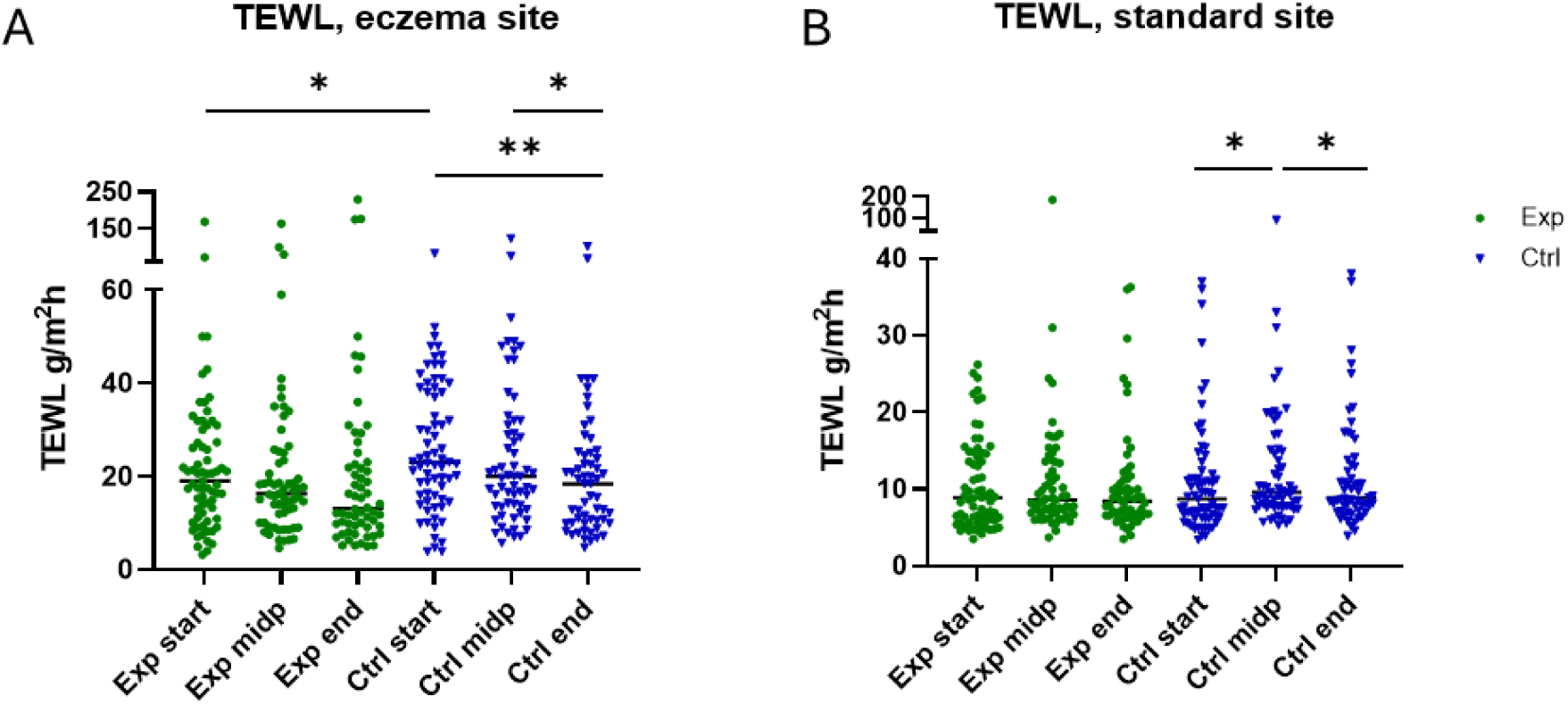
Changes in water evaporation through the skin. Transepidermal water loss (TEWL) at A) eczema and B) standard research areas. * p ≤ 0.05, ** p ≤ 0.01. Exp = Experimental group, Ctrl = control group. Lines represent medians.

At the standard research area TEWL value stayed constant over the study period in the experimental group (Fig 8B, medians 8.85, 8.6, 8.4), while the TEWL increased in the control group in winter (Fig 8B, medians 8.7, 9.6, 8.85, Supplementary Table 2B). In women, TEWL of standard research area was at significantly lower level (p=0.034) in experimental group in midpoint compared to control (medians 7.7. in exp and 10.1 in ctrl group) without difference at the start (p=0.340).

### Microbial extract prevented the worsening of erythema of the eczema during winter

Erythema in eczema site worsened in the control group in winter (Fig 9A, Supplementary Table 3A). The increase was significant from start to midpoint (p=0.0063) after which it decreased (p=0.0281) towards the end of the study. At the standard research site, the seasonal change was equal in both study groups (Fig 9B, descriptive statistics in Supplementary data Table 3B). Erythema was higher both in the midpoint and end of the study as compared to the start (p-values <0.001 from start to midpoint and from start to end in both groups).

**Figure 9.**
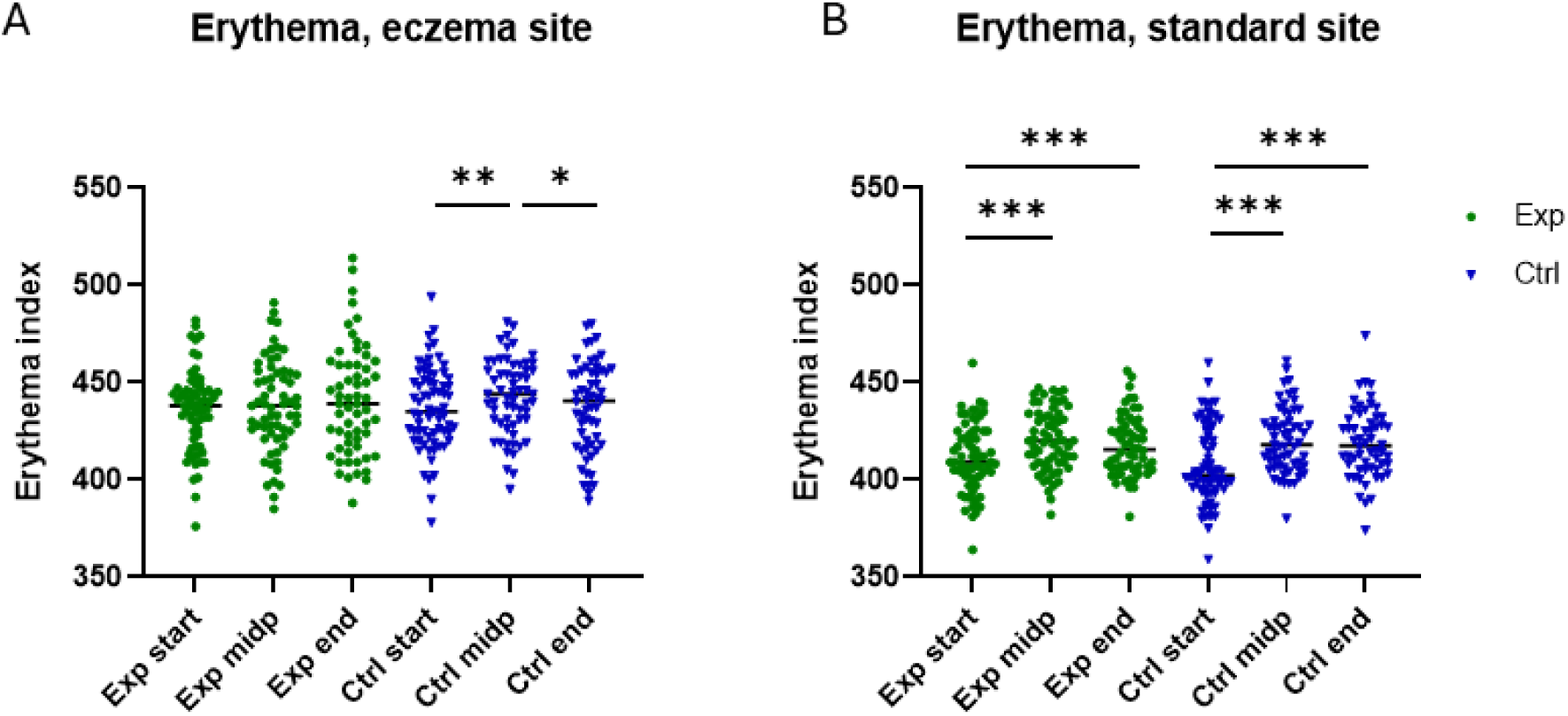
Changes in skin erythema index at A) eczema and B) standard research areas. * p ≤ 0.05, ** p ≤ 0.01. *** p≤ 0.001 Exp = Experimental group, Ctrl = control group. Lines represent means.

#### There were no differences in skin pH, antihistamine use and NRS itch between the trial arms

Skin pH value in eczema research area increased in both groups during the study (Supplementary Figure 6A). Skin pH value in standard research area was lower than in eczema area in every time points (start p<0.001, midpoint p=0.025, end p=0.031). There were no differences in skin pH values between the groups in either of the time points. Skin pH value in standard research area increased in both groups during the study (Supplementary Figure 6B). In the experimental group there was an increase from start to midpoint (mean increased from 5.12 to 5.26, p=0.02) and in control group there was increase from start to end (means 5.12 and 5.31, p=0.005) and from midpoint to end (means 5.19 and 5.31, p=0.008).

There were no differences between the groups in antihistamine use during the study. In the experimental group participants used antihistamine on average 19.7% of total days throughout the study and participants in control group used antihistamine 20.8% of total days. Some participants used antihistamine routinely on daily basis regardless of their symptoms to prevent future symptoms, which could affect the results. Allergy symptoms were not systemically recorded, but some participants reported reduced allergy symptoms for food (fish, milk), pollen or pets. There were 8 such reported cases in the experimental group and 2 cases in control group (p=0.097).

NRS itch question was included in the biweekly questionnaire from 1^st^ November to the end of the trial. There were no differences between the groups in NRS itch in any of the questionnaires, overall medians were 3.90 in experimental group and 3.84 in control group (Supplementary Fig 7).

### No differences in the test lotion dosage or in skin related adverse events between the trial arms

On average, the provided skin lotion was used 4.64 days per week in experimental group and 4.59 days per week in control group. There was no difference in application frequence between the groups or between the genders. The lotion was advised to be applied at least on arms and designated eczema area, but it was recommended to be applied also on other areas of the body to maximize exposure. This recommendation was well followed as on average 75 % of the participants in both groups applied test lotion on other areas than arms as well. The number of lotion bottles used throughout the study was on average 2.53 bottles (1265 g) in the experimental group and 2.63 bottles (1315 g) in the control group.

Participants were asked to report biweekly skin related adverse events (temporary or continuous itching, feeling of heat, redness, drying of the skin, urticaria, pustules) as well as other adverse events (free word) and events that have an effect on AD or are relevant for the study (skin related antibiotic use, use of clinical phototherapy or travels abroad, wherein there is more sunlight than in Finland during the study period). Also visits to dermatologist were asked. There were no significant differences between the groups in skin related adverse events, other adverse events, number of visits to dermatologist, skin related antibiotic use, phototherapy or travelling abroad (Supplementary Table 4).

### There were more correlations in the experimental groups between the variables

The selected physical measurements from the skin, TEWL and Erythema index, were good at indicating eczema severity, as they correlated with EASI in every visit (Supplementary table 5). Also, other results (NRS itch, POEM, medication days %) correlated with each other and with the physical measurements and EASI (Supplementary Fig 8). EASI of the last visit had negative correlation with application frequence of the test lotion (R_s_=-0.198, p=0.031) indicating that the more test lotion was used, the lower was EASI at the end of the study period. The test lotion application frequence was negatively correlated with POEM (R_s_=-0.176, p=0.038), indicating that the more test lotion was used, the less there were atopy symptoms. The benefit of frequent use of moisturizer is also seen in Fig 4.

Interestingly, when groups were analyzed separately, there were more correlations between the variables in the experimental group compared to control (Supplementary Fig 9). Medication days correlated with NRS itch (R_s_= 0.325 p=0.007), POEM (R_s_= 0.414, p<0.001) and EASI (by the midpoint R_s_= 0.420, p=0.001 and by the end R_s_= 0.409, p=0.001) in the experimental group while in the control group the medication days did not correlate with EASI (Supplementary Fig 9, 10). In the analysis of the background data of trial participants, a negative correlation was found between pollen allergy as an adult with the existence of a pet in a childhood family (R_s_=-0.236, p=0.015). There were no correlations between reported skin related adverse events and pollen or fungi allergies of the participants.

## Discussion

The present trial was the first randomized clinical trial to evaluate the effects of regular natural microbial exposure on the symptoms of AD in the context of the biodiversity hypothesis. Earlier studies indicate that exposure to environmental biodiversity may prevent atopic sensitization, but there are no results on the effect of biodiversity intervention on an already existing disease. The intervention, carried out using a skin lotion containing inactivated microbial extract, clearly decreased the need of atopy medications suggesting that it had a beneficial effect on AD. The potential explanations include improved skin barrier function leading to decreased erythema and irritation, and improved immune regulation as seen in our previous nature-based interventions ^18,19^. Our earlier preclinical trials have shown that exposure to inactivated bacteria has this kind of immunological effects in mice ^58,59^. The current study is the first to show that exposure to inactivated microbial diversity could also be beneficial in the treatment of atopic diseases in humans. This also in line with our previous study ^35^ which showed that combined use of corticosteroid crème and skin lotion containing inactivated microbial extract was more efficient in reducing pro-inflammatory cytokine levels in skin compared to corticosteroid + placebo (the lotion without microbes).

Inactivated microbial extract as the exposure material was well tolerated as there were no differences in adverse events between the experimental and control groups. For example, previous pollen allergy did not correlate with the reported adverse events, although the microbial extract was plant based.

We observed more correlation between the variables measured within the experimental group which provides further evidence for the biological response to treatment. Meaning that the members of the experimental group became more homogenous during the study.

Since the innate immune system responds to the microbial exposure of the environment and modulates the development of atopic diseases, it is tempting to speculate on the possible mechanisms behind our findings. Epithelial innate immune responses are not just inflammatory, but they also enhance cutaneous physical, chemical, microbial and immunological barriers ^60^. Highly diverse microbial extract contains components from gram negative and positive bacteria potentially including proteins, nucleic acids, carbohydrates, lipids and metabolites, and therefore it activates pattern recognition receptors on keratinocytes and immune cells in the skin. Toll-like receptors (TLR), one class of pattern-recognition receptors, are major initiators of innate immune responses and they contribute to the maintenance of tissue integrity in many sites of host-microbial interface ^61–65^. It is known that activation of toll-like receptor 2 enhances skin barrier function in humans ^66^ and enhances tight junction barrier in epidermal keratinocytes in vitro ^67,68^. TLRs have role both in stimulatory and regulatory arms of the immune response ^69–71^ and enhanced regulation downregulates pro-inflammatory cytokines improving skin barrier function ^36^. In a previous study we observed a downregulation of cytokines related to Th2, Th17 and Th22 cell subsets in atopic skin after four weeks use of microbial extract in lotion, but as a small pilot trial, this study was not able to distinguish the effect of microbial extract from the placebo (the lotion itself) ^35^. Contact to natural microbial diversity and microbial particles has been related to increased immunoregulation ^18,19,70^, and repeated exposure to microbial particles can also create immunotolerance that may help with chronic inflammation and autoimmune diseases ^72–74^. Microbially activated Th1 responses also counterbalance Th2 type allergic inflammation ^72,75,76^.

Many participants reported that they used the test lotion more regularly than they usually use moisturizers, and therefore both groups probably benefited from the moisturizing effect of the increased lotion use, as seen also in decreased need of medication (Fig 4). This was supported by the observed negative correlation between the test lotion application frequency and EASI scores at the last visit. Skin pH values increased in both groups indicating that pH could be increased due to enhanced use of moisturizer ^77^, or it could also be related to wintertime as AD patients are more sensitive to changes in skin temperature, which can be seen as increased skin pH ^78^. The influence of cold Finnish winter on eczema was clear as both POEM and number of medicine users increased at the time point when thermal winter begun (between 15^th^ Oct – 15^th^ Nov, variance depends on location) and there were increase of TEWL and Erythema index in winter, especially at the control group.

Our finding that a pet in childhood family was negatively correlated to pollen allergy is in line with earlier studies. It is discovered that contact with dogs in the first year of life as well as to furred or feathery pet in the first two years of life are negatively correlated to pollen sensitization ^79,80^, atopic sensitization, allergy and asthma ^81–85^.

We detected a potential influence of sex on the efficacy of microbial extract – women showed more clear reduction in topical corticosteroids and other AD treatments. It is known that women have more autoimmune diseases and AD ^86,87^ and many immunological responses may differ between sexes, like reduced response to glucocorticoids in women due to progesterone ^88^. The test lotion application frequency or baseline eczema severity did not differ between men and women. Nonetheless, the significantly lower number of male participants compared to females (29 vs 110) makes it difficult to compare the efficacy of this intervention in males and females.

This study has some limitations which need to be considered with interpreting the results. The aim of the trial was to study the effect of regular use of lotion containing inactivated microbial extract on AD skin. This study is, however, not a medical trial and the use of AD medication was not limited during the study (except the use of those medicine listed in the exclusion criteria) rendering medication as a confounding factor. Most of the trial participants were women, and the group of men was clearly smaller. In addition, the study participants had mainly Finnish genetic makeup, and therefore further studies are needed to confirm the effect in other populations. Finland has also a clear winter season when AD usually becomes more severe and when the main beneficial effect of the current intervention was seen. Therefore, it would be good to carry out similar trials in more temperate climates. One additional topic for further research is the dose in the extract – the present trial tested only one dose of the extract, and it is possible that a stronger beneficial effect could have been obtained with a higher dose.

Altogether, the results of this trial suggest that delivering inactivated microbial extract as nature exposure material in a skin lotion could be a beneficial addition to the daily care of AD. The purpose of the microbial extract is to provide regular contact with microbial diversity derived from natural materials, which differs it from the already established bacteriotherapy that aim at supporting skin microbiome ^47,89^. Current AD medicines focus on immunosuppression which is related to concern of side effects ^44^. Allergen-specific immunotherapy has been in use already a century for restoring normal immunity against certain allergens by creating peripheral tolerance to specific allergen ^90^, and novel therapies use innate immunity activators to induce tolerance against allergens ^91^.

We conclude that nature exposure by regular use of inactivated microbial extract in lotion was able to prevent worsening of the AD skin in winter. Importantly, the treatment reduced the use of atopy medication compared to placebo, despite the fact that AD severity was similar between the groups. The inactivated microbial extract as nature exposure material was also well tolerated. These results open new insights for AD treatment and underline the importance of exposure to natural microbial diversity in daily life. The results of this study encourage us also to carry out similar studies in other immune-mediated diseases.

## Data Availability

All data produced in the present study are available upon reasonable request to the authors

## Acknowledgements

We thank research nurse Saija Snygg for being an essential part of the research team. A special thank you is owed to the study participants for their participation in this study. The authors wish to acknowledge CSC – IT Center for Science, Finland, for computational resources.

## Author contributions

J.K. participated in the planning and implementation of the study, recruited the participants, handled the clinical visits, performed the statistical analyses, prepared the figures and tables, and wrote the manuscript. I.M. participated in the planning and implementation, handled the clinical visits, sent the bi-weekly questionnaires and participated in the result analysis and writing the manuscript. H.Hu. participated in the analysis of the results. H.Hy. participated in the planning and analysis of the results and writing the manuscript. V.H. participated in the analysis of the results and manuscript preparation. O.H.L. participated in the planning and the conduction of the trials, analysis of the results and manuscript preparation. A.S. participated in the planning of the trial and analysis of the results as well as manuscript preparation.

