## Supplementary material for "Microbial extract as a nature exposure material reduces the need for atopic dermatitis medication"

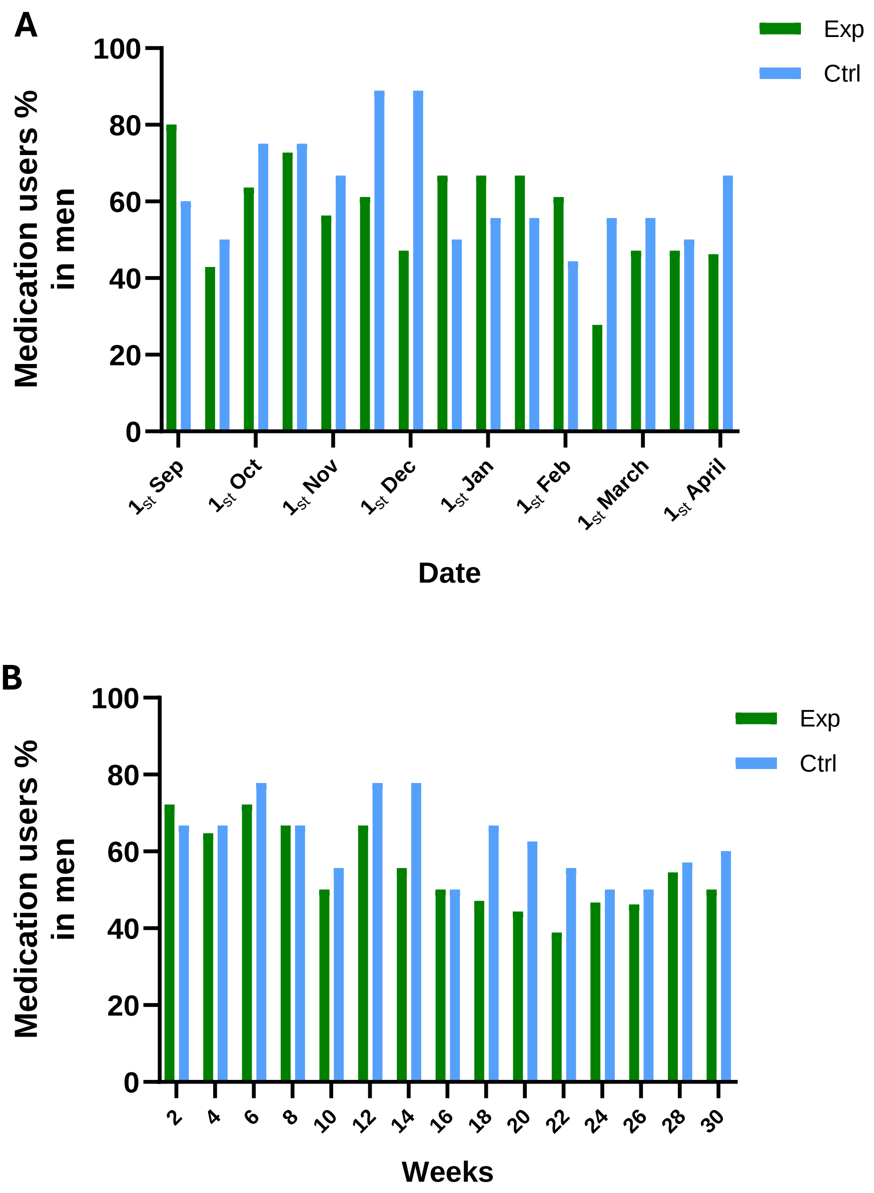

**Supplementary figure 1.** Medication users in men at A) chronologically and B) in weeks from the study start. Exp=experimental group, Ctrl=control group.

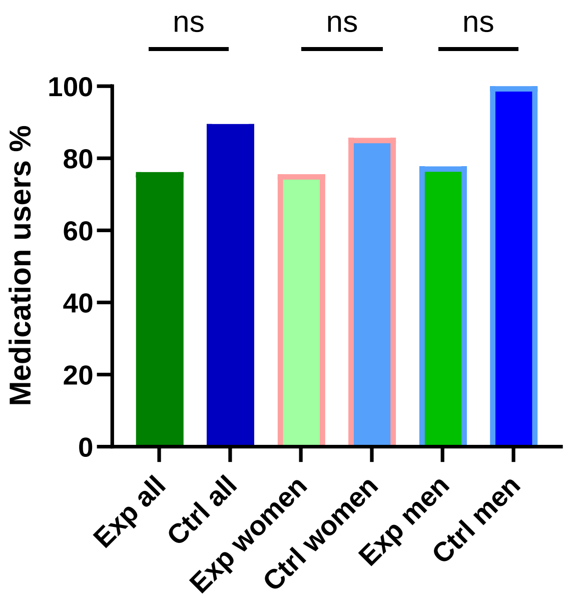

**Supplementary figure 2.** There were no differences in the number of AD medication users between the groups before the study. Results refer to three months' time and are collected from the preliminary interviews (0-60 days before the study start time). P-values were 0.091 (all participants), 0.662 (women) and 0.268 (men). Exp=experimental group, Ctrl=control group.

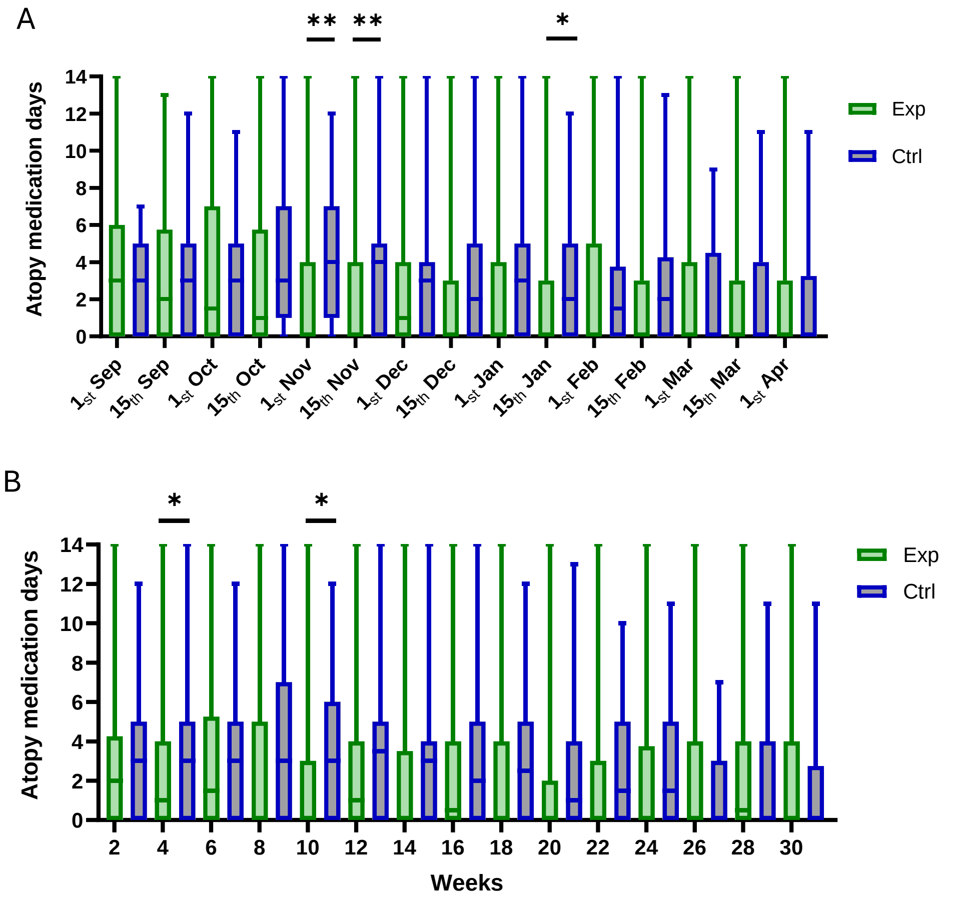

**Supplementary figure 3.** The experimental group (Exp) had less atopy medication days in several time points during the study compared to control group (Ctrl) when data is arranged A) chronologically and B) weeks from study start date. Median levels in experimental group are lower compared to control group in most of the time points. Whiskers represents range (min and max values), and boxes 2nd and 3rd quartiles with median line.

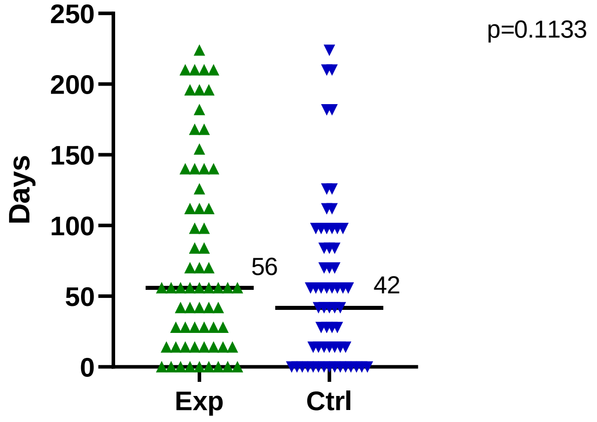

**Supplementary figure 4.** Comparison of the longest atopy medication free time between the groups. Lines represents medians. Exp=experimental group, Ctrl=control group.

**Supplementary table 1.** Descriptive statistics for Eczema area severity index (EASI).

| | Exp start | Exp midp | Exp end | Ctrl start | Ctrl midp | Ctrl end |
|---|---|---|---|---|---|---|
| Number of values | 70 | 64 | 60 | 69 | 61 | 59 |
| 25% percentile | 1.875 | 0.80 | 0.80 | 2.40 | 1.50 | 0.80 |
| Median | 4.65 | 2.55 | 2.85 | 5.1 | 3.2 | 1.7 |
| 75% percentile | 11.93 | 10.45 | 6.58 | 9.40 | 10.15 | 5.10 |

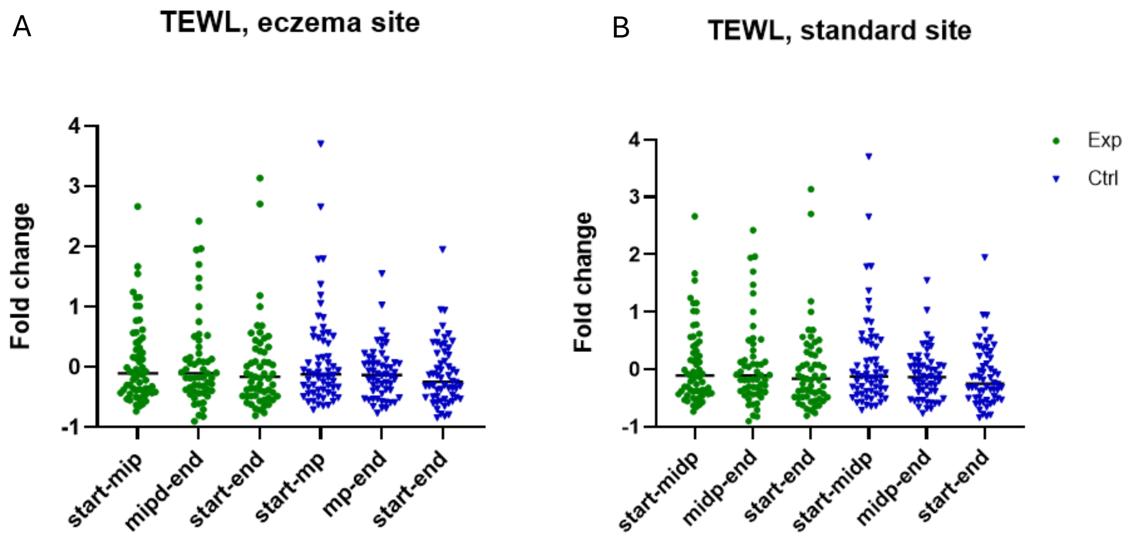

**Supplementary figure 5**. TEWL of eczema site presented as fold changes (the subsequent value divided by the original value), p-values were 0.950 (from start to midpoint), 0.359 (from midpoint to end) and 0.457 (from start to end), Exp= experimental group, Ctrl=control group.

**Supplementary table 2.** Descriptive statistics of the TEWL results A. in eczema and B. in standard research area. These tables refer to figures 8A and B in the article.

| A | Exp start | Exp midp | Exp end | Ctrl start | Ctrl midp | Ctrl end |
|---|---|---|---|---|---|---|
| Number of values | 69 | 63 | 60 | 69 | 61 | 58 |
| 25 % Percentile | 11.70 | 10.00 | 9.33 | 15.50 | 13.50 | 9.98 |
| **Median** | **19.00** | **16.30** | **13.15** | **23.00** | **20.00** | **18.40** |
| 75 % Percentile | 28.65 | 25.10 | 22.98 | 38.50 | 31.50 | 25.08 |

| B | Exp start | Exp midp | Exp end | Ctrl start | Ctrl midp | Ctrl end |
|---|---|---|---|---|---|---|
| Number of values | 70 | 63 | 60 | 69 | 61 | 58 |
| 25 % Percentile | 5.98 | 6.90 | 6.63 | 6.35 | 7.80 | 7.18 |
| **Median** | **8.85** | **8.60** | **8.40** | **8.70** | **9.60** | **8.85** |
| 75 % Percentile | 14.63 | 13.50 | 11.08 | 11.65 | 15.00 | 13.35 |

**Supplementary table 3.** Descriptive statistics of the Erythema index results A. in eczema and B. in standard research area. These tables refer to figures 9A and B in the article.

| A | Exp start | Exp midp | Exp end | Ctrl start | Ctrl midp | Ctrl end |
|---|---|---|---|---|---|---|
| Number of values | 70 | 64 | 60 | 69 | 61 | 58 |
| **Mean** | **435.8** | **438.1** | **440.7** | **436.7** | **442.5** | **436.7** |
| Std. Deviation | 20.38 | 23.69 | 28.15 | 21.21 | 19.43 | 23.37 |

| B | Exp start | Exp midp | Exp end | Ctrl start | Ctrl midp | Ctrl end |
|---|---|---|---|---|---|---|
| Number of values | 70 | 64 | 60 | 69 | 61 | 58 |
| **Mean** | **411.3** | **420.4** | **417.5** | **407.6** | **420.2** | **418.8** |
| Std. Deviation | 17.42 | 15.74 | 15.54 | 20.14 | 16.60 | 17.95 |

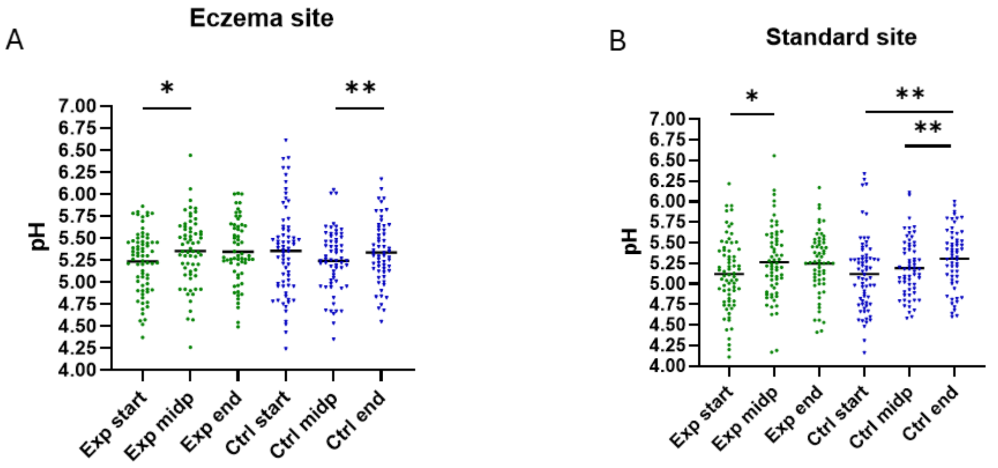

| | Exp start | Exp midp | Exp end | Ctrl start | Ctrl midp | Ctrl end |
|---|---|---|---|---|---|---|
| Number of values | 71 | 64 | 60 | 69 | 62 | 59 |
| Mean | 5.12 | 5.26 | 5.25 | 5.12 | 5.19 | 5.31 |
| Std. Deviation | 0.44 | 0.44 | 0.38 | 0.45 | 0.35 | 0.37 |

| | Exp start | Exp midp | Exp end | Ctrl start | Ctrl midp | Ctrl end |
|---|---|---|---|---|---|---|
| Number of values | 71 | 64 | 60 | 68 | 62 | 59 |
| Mean | 5.23 | 5.35 | 5.34 | 5.35 | 5.24 | 5.34 |
| Std. Deviation | 0.35 | 0.39 | 0.37 | 0.52 | 0.36 | 0.36 |

**Supplementary figure 6**. Skin pH value in A) eczema and B) standard research areas. Exp= experimental group, Ctrl=control group.

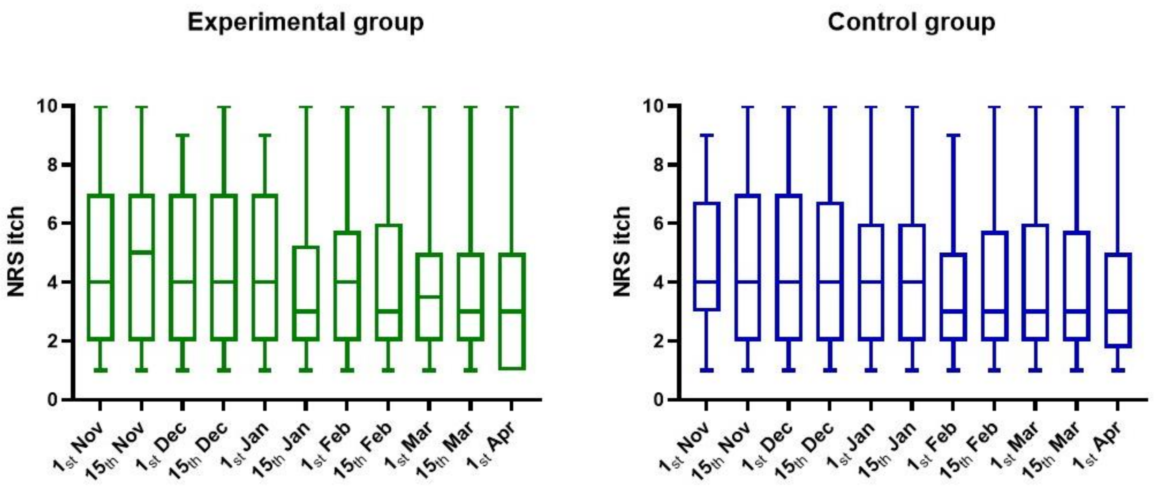

**Supplementary figure 7.** NRS itch results on each questionnaire in experimental and control groups. Data includes answers from all participants. Whiskers represents range (min and max values), and boxes 2nd and 3rd quartiles with median line.

**Supplementary table 4.** Reported skin related adverse events and miscellaneous events that are relevant for the study from biweekly questionnaires as percentages in each group. Percentages represent the proportion of participants that reported the named event during the study from total number of people in that group.

| **Reported skin related adverse events** | Experimental % | Control % |
|---|---|---|
| Temporary itching, feeling of heat, redness | 19 | 15 |
| Continuous itching, feeling of heat, redness | 10 | 3 |
| Drying of the skin | 4 | 8 |
| Urticaria | 3 | 7 |
| Pustules | 9 | 6 |
| **Other adverse events and events relevant for the study** | | |
| Other reported skin related adverse events | 21 | 13 |
| Adverse events reported at last visit | 6 | 7 |
| Antibiotic use where association to study cannot be excluded | 3 | 1 |
| Visits to dermatologist | 17 | 23 |
| Polyclinical photherapy | 1 | 4 |
| Travel abroad | 29 | 23 |

**Supplementary table 5.** Correlations between TEWL (transepidermal water loss) and Erythema index of the eczema research area and EASI (eczema area severity index). All participants are included in the analysis.

| **Time** | **Method** | **Erythema index** | **EASI** | **AD medication** |
|---|---|---|---|---|
| Start | **TEWL** | 0.528** (p<0.001) | 0.254** (p=0.003) | |
| | **Erythema index** | | 0.242** (p=0.004) | |
| Midpoint | **TEWL** | 0.534** (p<0.001) | 0.369** (p<0.001) | 0.212* (p=0.028) |
| | **Erythema index** | | 0.300** (p<0.001) | 0.228* (p=0.017) |
| | **EASI** | | | 0.463** (p<0.001) |
| End | **TEWL** | 0.677** (p<0.001) | 0.330** (p<0.001) | ns |
| | **Erythema index** | | 0,204* (p=0,027) | ns |
| | **EASI** | | | 0,334** (p<0,001) |

**Supplementary material** for the manuscript “Microbial extract as a nature exposure material reduces the need for atopic dermatitis medication” Kalmari et al. 2025

**Correlations**

| | | | Test lotion application frequency | TEWL_stand_midpoint | TEWL_stand_end | TEWL_eczema_midpoint | TEWL_eczema_end | Medication days % | NRS itch | POEM | Erythema_stand_midpoint | Erythema_stand_end | Erythema_ecz_midpoint | Erythema_eczema_end | pH_stand_midpoint | pH_stand_end | pH_ecz_midpoint | pH_ecz_end | EASI2 | EASI3 |
|---|---|---|---|---|---|---|---|---|---|---|---|---|---|---|---|---|---|---|---|---|
| Spearman's rho | TEWL_stand_midpoint | Correlation Coefficient | | | | | | | | | | | | | | | | | | |
| | | Sig. (2-tailed) | | | | | | | | | | | | | | | | | | |
| | | N | | | | | | | | | | | | | | | | | | |
| | TEWL_stand_end | Correlation Coefficient | | .705** | | | | | | | | | | | | | | | | |
| | | Sig. (2-tailed) | | 0.000 | | | | | | | | | | | | | | | | |
| | | N | | 113 | | | | | | | | | | | | | | | | |
| | TEWL_eczema_midpoint | Correlation Coefficient | | .468** | .357** | | | | | | | | | | | | | | | |
| | | Sig. (2-tailed) | | 0.000 | 0.000 | | | | | | | | | | | | | | | |
| | | N | | 124 | 113 | | | | | | | | | | | | | | | |
| | TEWL_eczema_end | Correlation Coefficient | | .309** | .538** | .604** | | | | | | | | | | | | | | |
| | | Sig. (2-tailed) | | 0.001 | 0.000 | 0.000 | | | | | | | | | | | | | | |
| | | N | | 113 | 118 | 113 | | | | | | | | | | | | | | |
| | Medication days % | Correlation Coefficient | | | | | | | | | | | | | | | | | | |
| | | Sig. (2-tailed) | | | | | | | | | | | | | | | | | | |
| | | N | | | | | | | | | | | | | | | | | | |
| | NRS itch | Correlation Coefficient | | .274** | .214* | | .241** | .261** | | | | | | | | | | | | |
| | | Sig. (2-tailed) | | 0.002 | 0.020 | | 0.008 | 0.002 | | | | | | | | | | | | |
| | | N | | 124 | 118 | | 118 | 137 | | | | | | | | | | | | |
| | POEM | Correlation Coefficient | -.176* | .235** | .324** | .257** | .314** | .247** | .645** | | | | | | | | | | | |
| | | Sig. (2-tailed) | 0.038 | 0.009 | 0.000 | 0.004 | 0.001 | 0.003 | 0.000 | | | | | | | | | | | |
| | | N | 139 | 124 | 118 | 124 | 118 | 139 | 137 | | | | | | | | | | | |
| | Erythema_stand_midpoint | Correlation Coefficient | | .390** | .246** | | .190* | | | | | | | | | | | | | |
| | | Sig. (2-tailed) | | 0.000 | 0.008 | | 0.043 | | | | | | | | | | | | | |
| | | N | | 124 | 114 | | 114 | | | | | | | | | | | | | |
| | Erythema_stand_end | Correlation Coefficient | | .310** | .254** | | .229* | | .181* | | .778** | | | | | | | | | |
| | | Sig. (2-tailed) | | 0.001 | 0.005 | | 0.013 | | 0.049 | | 0.000 | | | | | | | | | |
| | | N | | 113 | 118 | | 118 | | 118 | | 114 | | | | | | | | | |
| | Erythema_ecz_midpoint | Correlation Coefficient | | .208* | .245** | .534** | .553** | .185* | | | .441** | .339** | | | | | | | | |
| | | Sig. (2-tailed) | | 0.021 | 0.009 | 0.000 | 0.000 | 0.038 | | | 0.000 | 0.000 | | | | | | | | |
| | | N | | 124 | 114 | 124 | 114 | 125 | | | 125 | 114 | | | | | | | | |
| | Erythema_eczema_end | Correlation Coefficient | | | .222* | .464** | .677** | | | .194* | .306** | .403** | .697** | | | | | | | |
| | | Sig. (2-tailed) | | | 0.0156 | 0.000 | 0.000 | | | 0.035 | 0.001 | 0.000 | 0.000 | | | | | | | |
| | | N | | | 118 | 113 | 118 | | | 118 | 114 | 118 | 114 | | | | | | | |
| | pH_stand_midpoint | Correlation Coefficient | | | | | | | | | | | | | | | | | | |
| | | Sig. (2-tailed) | | | | | | | | | | | | | | | | | | |
| | | N | | | | | | | | | | | | | | | | | | |
| | pH_stand_end | Correlation Coefficient | | | | | | | | | | | | | .471** | | | | | |
| | | Sig. (2-tailed) | | | | | | | | | | | | | 0.000 | | | | | |
| | | N | | | | | | | | | | | | | 113 | | | | | |
| | pH_ecz_midpoint | Correlation Coefficient | | | | | | | | | | | | | .576** | .355** | | | | |
| | | Sig. (2-tailed) | | | | | | | | | | | | | 0.000 | 0.000 | | | | |
| | | N | | | | | | | | | | | | | 124 | 113 | | | | |
| | pH_eczema_end | Correlation Coefficient | | | | | | | | | | | | | .384** | .629** | .448** | | | |
| | | Sig. (2-tailed) | | | | | | | | | | | | | 0.000 | 0.000 | 0.000 | | | |
| | | N | | | | | | | | | | | | | 113 | 118 | 113 | | | |
| | EASI2 | Correlation Coefficient | | .457** | .352** | .369** | .314** | .320** | .535** | .464** | .287** | .252** | .300** | .261** | | | | | | |
| | | Sig. (2-tailed) | | 0.000 | 0.000 | 0.000 | 0.001 | 0.000 | 0.000 | 0.000 | 0.001 | 0.007 | 0.001 | 0.005 | | | | | | |
| | | N | | 124 | 114 | 124 | 114 | 125 | 125 | 125 | 125 | 114 | 125 | 114 | | | | | | |
| | EASI3 | Correlation Coefficient | -.198* | .365** | .378** | .295** | .330** | .298** | .599** | .598** | .224* | .274** | .199* | .204* | | | | | .734** | |
| | | Sig. (2-tailed) | 0.031 | 0.000 | 0.000 | 0.001 | 0.000 | 0.001 | 0.000 | 0.000 | 0.016 | 0.003 | 0.033 | 0.027 | | | | | 0.000 | |
| | | N | 119 | 114 | 118 | 114 | 118 | 119 | 119 | 119 | 115 | 118 | 115 | 118 | | | | | 115 | |

*. Correlation is significant at the 0.05 level (2-tailed).

**. Correlation is significant at the 0.01 level (2-tailed).

**Supplementary figure 8.** Significant correlations between the methods when both groups are combined. Results are from clinical visits (TEWL, Erythema index, pH, EASI) and from biweekly questionnaires (test lotion application frequence, NRS itch, POEM, medication days %).

**Supplementary material** for the manuscript “Microbial extract as a nature exposure material reduces the need for atopic dermatitis medication” Kalmari et al. 2025

**A** **EXP-group**

| | | | Test lotion application | TEWL_stand_midpoint | TEWL_stand_end | TEWL_eczema_midpoint | TEWL_eczema_end | Medication days % | NRS itch | POEM | Erythema_stand_midpoint | Erythema_stand_end | Erythema_ecz_midpoint | Erythema_eczema_end | pH_stand_midpoint | pH_stand_end | pH_ecz_midpoint | pH_ecz_end | EASI2 | EASI3 |
|---|---|---|---|---|---|---|---|---|---|---|---|---|---|---|---|---|---|---|---|---|
| Spearman's rho | TEWL_stand_midpoint | Correlation Coefficient | | | | | | | | | | | | | | | | | | |
| | | Sig. (2-tailed) | | | | | | | | | | | | | | | | | | |
| | | N | | | | | | | | | | | | | | | | | | |
| | TEWL_stand_end | Correlation Coefficient | | .740** | | | | | | | | | | | | | | | | |
| | | Sig. (2-tailed) | | 0.000 | | | | | | | | | | | | | | | | |
| | | N | | 57 | | | | | | | | | | | | | | | | |
| | TEWL_eczema_midpoint | Correlation Coefficient | | .449** | .318* | | | | | | | | | | | | | | | |
| | | Sig. (2-tailed) | | 0.000 | 0.016 | | | | | | | | | | | | | | | |
| | | N | | 63 | 57 | | | | | | | | | | | | | | | |
| | TEWL_eczema_end | Correlation Coefficient | | .343** | .494** | .553** | | | | | | | | | | | | | | |
| | | Sig. (2-tailed) | | 0.009 | 0.000 | 0.000 | | | | | | | | | | | | | | |
| | | N | | 57 | 60 | 57 | | | | | | | | | | | | | | |
| | Medication days % | Correlation Coefficient | | | | | | | | | | | | | | | | | | |
| | | Sig. (2-tailed) | | | | | | | | | | | | | | | | | | |
| | | N | | | | | | | | | | | | | | | | | | |
| | NRS itch | Correlation Coefficient | | .285* | | | | .325** | | | | | | | | | | | | |
| | | Sig. (2-tailed) | | 0.024 | | | | 0.007 | | | | | | | | | | | | |
| | | N | | 63 | | | | 68 | | | | | | | | | | | | |
| | POEM | Correlation Coefficient | | | .316* | | .300* | .414** | .665** | | | | | | | | | | | |
| | | Sig. (2-tailed) | | | 0.014 | | 0.020 | 0.000 | 0.000 | | | | | | | | | | | |
| | | N | | | 60 | | 60 | 70 | 68 | | | | | | | | | | | |
| | Erythema_stand_midpoint | Correlation Coefficient | | .433** | | | .331* | | | | | | | | | | | | | |
| | | Sig. (2-tailed) | | 0.000 | | | 0.011 | | | | | | | | | | | | | |
| | | N | | 63 | | | 58 | | | | | | | | | | | | | |
| | Erythema_stand_end | Correlation Coefficient | | .274* | | | .358** | | | | .792** | | | | | | | | | |
| | | Sig. (2-tailed) | | 0.039 | | | 0.005 | | | | 0.000 | | | | | | | | | |
| | | N | | 57 | | | 60 | | | | 58 | | | | | | | | | |
| | Erythema_ecz_midpoint | Correlation Coefficient | | .292* | | .578** | .638** | | | | .470** | .456** | | | | | | | | |
| | | Sig. (2-tailed) | | 0.020 | | 0.000 | 0.000 | | | | 0.000 | 0.000 | | | | | | | | |
| | | N | | 63 | | 63 | 58 | | | | 64 | 58 | | | | | | | | |
| | Erythema_eczema_end | Correlation Coefficient | | | .328* | .526** | .816** | | | | .360** | .495** | .764** | | | | | | | |
| | | Sig. (2-tailed) | | | 0.011 | 0.000 | 0.000 | | | | 0.005 | 0.000 | 0.000 | | | | | | | |
| | | N | | | 60 | 57 | 60 | | | | 58 | 60 | 58 | | | | | | | |
| | pH_stand_midpoint | Correlation Coefficient | | | | | | | | | | | | | | | | | | |
| | | Sig. (2-tailed) | | | | | | | | | | | | | | | | | | |
| | | N | | | | | | | | | | | | | | | | | | |
| | pH_stand_end | Correlation Coefficient | | | | | | | | | | | | | .470** | | | | | |
| | | Sig. (2-tailed) | | | | | | | | | | | | | 0.000 | | | | | |
| | | N | | | | | | | | | | | | | 57 | | | | | |
| | pH_ecz_midpoint | Correlation Coefficient | | | | | | | | | | | | | .649** | .433** | | | | |
| | | Sig. (2-tailed) | | | | | | | | | | | | | 0.000 | 0.001 | | | | |
| | | N | | | | | | | | | | | | | 63 | 57 | | | | |
| | pH_eczema_end | Correlation Coefficient | | | | -.279* | | | | | | .269* | | | .406** | .639** | .373** | | | |
| | | Sig. (2-tailed) | | | | 0.035 | | | | | | 0.038 | | | 0.002 | 0.000 | 0.004 | | | |
| | | N | | | | 57 | | | | | | 60 | | | 57 | 60 | 57 | | | |
| | EASI2 | Correlation Coefficient | | .480** | .347** | .378** | .364** | .430** | .580** | .515** | .296* | | .376** | .324* | | | | | | |
| | | Sig. (2-tailed) | | 0.000 | 0.008 | 0.002 | 0.005 | 0.000 | 0.000 | 0.000 | 0.017 | | 0.002 | 0.013 | | | | | | |
| | | N | | 63 | 58 | 63 | 58 | 64 | 64 | 64 | 64 | | 64 | 58 | | | | | | |
| | EASI3 | Correlation Coefficient | | .335* | .313* | .299* | .357** | .334** | .667** | .719** | | | | | | | | | .771** | |
| | | Sig. (2-tailed) | | 0.011 | 0.015 | 0.024 | 0.005 | 0.009 | 0.000 | 0.000 | | | | | | | | | 0.000 | |
| | | N | | 57 | 60 | 57 | 60 | 60 | 60 | 60 | | | | | | | | | 58 | |

**. Correlation is significant at the 0.01 level (2-tailed).
*. Correlation is significant at the 0.05 level (2-tailed).

**B** **CTRL-group**

| | | | Test lotion application | TEWL_stand_midpoint | TEWL_stand_end | TEWL_eczema_midpoint | TEWL_eczema_end | Medication days % | NRS itch | POEM | Erythema_stand_midpoint | Erythema_stand_end | Erythema_ecz_midpoint | Erythema_eczema_end | pH_stand_midpoint | pH_stand_end | pH_ecz_midpoint | pH_ecz_end | EASI2 | EASI3 |
|---|---|---|---|---|---|---|---|---|---|---|---|---|---|---|---|---|---|---|---|---|
| Spearman's rho | TEWL_stand_midpoint | Correlation Coefficient | | | | | | | | | | | | | | | | | | |
| | | Sig. (2-tailed) | | | | | | | | | | | | | | | | | | |
| | | N | | | | | | | | | | | | | | | | | | |
| | TEWL_stand_end | Correlation Coefficient | | .642** | | | | | | | | | | | | | | | | |
| | | Sig. (2-tailed) | | 0.000 | | | | | | | | | | | | | | | | |
| | | N | | 56 | | | | | | | | | | | | | | | | |
| | TEWL_eczema_midpoint | Correlation Coefficient | | .457** | .362** | | | | | | | | | | | | | | | |
| | | Sig. (2-tailed) | | 0.000 | 0.006 | | | | | | | | | | | | | | | |
| | | N | | 61 | 56 | | | | | | | | | | | | | | | |
| | TEWL_eczema_end | Correlation Coefficient | | .271* | .586** | .660** | | | | | | | | | | | | | | |
| | | Sig. (2-tailed) | | 0.043 | 0.000 | 0.000 | | | | | | | | | | | | | | |
| | | N | | 56 | 58 | 56 | | | | | | | | | | | | | | |
| | Medication days % | Correlation Coefficient | | | | | | | | | | | | | | | | | | |
| | | Sig. (2-tailed) | | | | | | | | | | | | | | | | | | |
| | | N | | | | | | | | | | | | | | | | | | |
| | NRS itch | Correlation Coefficient | | .256* | | | | | | | | | | | | | | | | |
| | | Sig. (2-tailed) | | 0.047 | | | | | | | | | | | | | | | | |
| | | N | | 61 | | | | | | | | | | | | | | | | |
| | POEM | Correlation Coefficient | | | .337** | .295* | .302* | | .615** | | | | | | | | | | | |
| | | Sig. (2-tailed) | | | 0.010 | 0.021 | 0.021 | | 0.000 | | | | | | | | | | | |
| | | N | | | 58 | 61 | 58 | | 69 | | | | | | | | | | | |
| | Erythema_stand_midpoint | Correlation Coefficient | | .363** | | | | | | | | | | | | | | | | |
| | | Sig. (2-tailed) | | 0.004 | | | | | | | | | | | | | | | | |
| | | N | | 61 | | | | | | | | | | | | | | | | |
| | Erythema_stand_end | Correlation Coefficient | | .324* | | | | | | | .761** | | | | | | | | | |
| | | Sig. (2-tailed) | | 0.015 | | | | | | | 0.000 | | | | | | | | | |
| | | N | | 56 | | | | | | | 56 | | | | | | | | | |
| | Erythema_ecz_midpoint | Correlation Coefficient | | | | .507** | .489** | | | | .433** | | | | | | | | | |
| | | Sig. (2-tailed) | | | | 0.000 | 0.000 | | | | 0.000 | | | | | | | | | |
| | | N | | | | 61 | 56 | | | | 61 | | | | | | | | | |
| | Erythema_eczema_end | Correlation Coefficient | | | | .427** | .523** | | | | | .302* | .637** | | | | | | | |
| | | Sig. (2-tailed) | | | | 0.001 | 0.000 | | | | | 0.021 | 0.000 | | | | | | | |
| | | N | | | | 56 | 58 | | | | | 58 | 56 | | | | | | | |
| | pH_stand_midpoint | Correlation Coefficient | | | | | | | | | | | | | | | | | | |
| | | Sig. (2-tailed) | | | | | | | | | | | | | | | | | | |
| | | N | | | | | | | | | | | | | | | | | | |
| | pH_stand_end | Correlation Coefficient | | | | | | | | | | | | | .490** | | | | | |
| | | Sig. (2-tailed) | | | | | | | | | | | | | 0.000 | | | | | |
| | | N | | | | | | | | | | | | | 56 | | | | | |
| | pH_ecz_midpoint | Correlation Coefficient | | | | | | | | | | | | | .491** | .294* | | | | |
| | | Sig. (2-tailed) | | | | | | | | | | | | | 0.000 | 0.028 | | | | |
| | | N | | | | | | | | | | | | | 61 | 56 | | | | |
| | pH_eczema_end | Correlation Coefficient | -.264* | | | | | | | | | | | | .347** | .615** | .519** | | | |
| | | Sig. (2-tailed) | 0.045 | | | | | | | | | | | | 0.009 | 0.000 | 0.000 | | | |
| | | N | 58 | | | | | | | | | | | | 56 | 58 | 56 | | | |
| | EASI2 | Correlation Coefficient | | .429** | .335* | .361** | | | .479** | .410** | .262* | | | | | | | | | |
| | | Sig. (2-tailed) | | 0.001 | 0.012 | 0.004 | | | 0.000 | 0.001 | 0.042 | | | | | | | | | |
| | | N | | 61 | 56 | 61 | | | 61 | 61 | 61 | | | | | | | | | |
| | EASI3 | Correlation Coefficient | | .400** | .450** | .282* | .285* | .263* | .525** | .458** | | .272* | | | | | | | .676** | |
| | | Sig. (2-tailed) | | 0.002 | 0.000 | 0.034 | 0.030 | 0.045 | 0.000 | 0.000 | | 0.039 | | | | | | | 0.000 | |
| | | N | | 57 | 58 | 57 | 58 | 59 | 59 | 59 | | 58 | | | | | | | 57 | |

*. Correlation is significant at the 0.05 level (2-tailed).
**. Correlation is significant at the 0.01 level (2-tailed).

**Supplementary figure 9.** Significant correlations in A) Experimental (Exp) and B) Control (Ctrl) groups separately. Correlations marked in green color are found in both groups and those with blue color are found only in this group. Results are from clinical visits (TEWL, Erythema index, pH, EASI) and from biweekly questionnaires (test lotion application frequence, NRS itch, POEM, medication days %).

**EXP**

**Correlations**

| | | | EASI2 | EASI3 |
|---|---|---|---|---|
| Spearman's rho | Med days_start to midpoint | Correlation Coefficient | 0.420** | |
| | | Sig. (2-tailed) | 0.001 | |
| | | N | 64 | |
| | Med days_midpoint to end | Correlation Coefficient | | 0.409** |
| | | Sig. (2-tailed) | | 0.001 |
| | | N | | 60 |

**. Correlation is significant at the 0.01 level (2-tailed).

**CTRL**

**Correlations**

| | | | EASI2 | EASI3 |
|---|---|---|---|---|
| Spearman's rho | Med days_start to midpoint | Correlation Coefficient | 0.209 | |
| | | Sig. (2-tailed) | 0.106 | |
| | | N | 61 | |
| | Med days_midpoint to end | Correlation Coefficient | | 0.128 |
| | | Sig. (2-tailed) | | 0.336 |
| | | N | | 59 |

**Supplementary figure 10.** Correlation of atopy medication days (%) before each clinical visit (from start to midpoint and from midpoint to end) with EASI scoring of midpoint (EASI2) and end (EASI3). There were correlations only in Experimental (Exp) group but not in Control (Ctrl) group.
